# RAPID: A Targeted Long-Read RNA Workflow for Functional Resolution of Splicing Variants in Rare Disease

**DOI:** 10.64898/2025.12.30.25342835

**Authors:** Kylie Montgomery, Hannah Macpherson, Claire Anderson, Charles Wade, Emil K Gustavsson, David S Lynch, Louise C Wilson, James Davison, Emma Wakeling, Karin Tuschl, Henry Houlden, Emma Clement, Phillipa Mills, Mina Ryten

## Abstract

**Background:** Molecular diagnosis of rare disease plateaus at ∼50%, partly due to technical limitations of short-read sequencing and the persistent challenge of interpreting variants of uncertain significance (VUS). Splice-altering variation represents a major source of unresolved cases, yet functional assessment remains difficult in routine practice.

**Methods:** We developed a fully modular, sample-to-answer workflow for targeted long-read RNA sequencing (lrRNA-seq) using Oxford Nanopore Technologies and applied it to six unsolved cases with suspected monogenic neurometabolic disease. Candidates were selected after WES/WGS and multidisciplinary team review (MDT) indicating ≤5 genes of interest. The workflow was designed to be diagnostically deployable, enabling near-full-length transcript assessment from accessible tissues without reliance on large control cohorts.

**Results:** lrRNA-seq yielded actionable findings for all six probands. It confirmed pathogenic splice disruption in two cases, prompted gene exclusion in one case, and generated RNA-level evidence prioritising further DNA investigation in three cases. Across these scenarios, lrRNA-seq provided direct, mechanism-level insight that either resolved diagnosis or refined variant interpretation. The workflow provided near-full-length isoform structures with reproducible single-sample interpretation and produced informative results within two working days at <£500 per sample reagent cost.

**Conclusion:** Targeted lrRNA-seq offers rapid, cost-effective functional evidence to resolve VUS, direct DNA follow-up, and support timely diagnosis in rare disease. The RAPID workflow demonstrates that long-read RNA sequencing can be implemented within existing diagnostic infrastructure and provides a scalable route to routine transcript-level assessment in clinical genomics.

## Main text

Rare monogenic disorders collectively affect hundreds of millions of people worldwide, with a disproportionate impact on infants and children.^1,2^ For affected families, securing a molecular diagnosis is not merely confirmatory: it guides disease-specific management, informs prognosis, enables reproductive planning, and increasingly determines access to targeted therapies and clinical trials. Despite widespread adoption of genome and exome sequencing, many individuals remain undiagnosed, often because variants are classified as variants of uncertain significance (VUS), which limits clinical actionability.^3^

A major contributor to this diagnostic gap is the set of non-coding variants that disrupt splicing.^4^ Frontline DNA-based tests prioritise coding regions and canonical splice sites, yet pathogenic mechanisms often arise from alternative splice junctions, cryptic exons, isoform usage, and regulatory variation outside these boundaries. In the absence of direct transcript evidence, predicted splicing effects remain inferential, and “single-hit” findings in recessive genes frequently lack demonstrable mechanisms on the second allele. Up to 62% of pathogenic single-nucleotide variants are estimated to affect splicing,^3^ underscoring the value of functional, transcript-guided follow-up in uncovering missed splice or regulatory defects.

Although tissue specificity can limit the utility of RNA-based diagnostics, many clinically relevant genes are nonetheless robustly expressed in readily accessible tissues. The greater challenge has been interpretability.^4^ Conventional short-read RNA-seq, while transformative for discovery, cannot faithfully reconstruct full-length transcripts and instead infers structure from short, fragmented alignments. This fragmented view complicates the resolution of exon connectivity, cryptic junctions, or alternative transcript usage, particularly in repetitive or incompletely annotated regions. As a result, distinguishing pathogenic events from normal isoform diversity typically requires comparison with additional samples processed in parallel. In diagnostic settings, however, true baseline controls are rarely available, leading laboratories to consider using other affected individuals within the same sequencing run as comparators, a practice that risks obscuring disease-relevant splicing changes and enforces batching of samples,^5,6^ thereby delaying analysis for individual patients.^7^

By enabling complete reconstruction of full-length transcripts, targeted long-read RNA sequencing overcomes the interpretative constraints that necessitate batching of controls in short-read workflows and makes transcript-level assessment far more practical in clinical settings. In this study, we describe a complete, modular, sample-to-result workflow for targeted long-read RNA sequencing, designed for translational use in diagnostic laboratories. We applied this approach to six unsolved rare-disease cases and demonstrate a threefold impact: (i) confirming the mechanistic consequences of splice-altering variants; (ii) quantifying transcript imbalance and assessing partial (“leaky”) splicing in accessible tissues relative to pooled controls to guide targeted DNA follow-up; and (iii) identifying transcript-level deviations that pinpoint regions of interest for second-allele searches in recessive disease.

To evaluate the clinical applicability of this approach, we focused on developing a workflow that mirrored the constraints and realities of diagnostic practice. This required selecting cases where transcript-level evidence could have immediate clinical impact, implementing laboratory protocols already familiar to diagnostic laboratories, and designing an analytical pipeline that was modular, reproducible, and sufficiently streamlined for routine use. We therefore undertook a pilot study of six unsolved rare-disease cases drawn from a previously sequenced cohort, applying targeted lrRNA-seq within a framework that enabled direct and clinically relevant assessment of its utility.

Six individuals meeting these criteria were selected from the Study of Inherited Metabolic Diseases (SIMD) and the International Genomics Collaboration (IGC) cohorts (see supplementary methods for further details), all of whom had previously undergone short-read WES or WGS and remained without a resolved molecular diagnosis. The cohort comprised individuals with rare monogenic neurogenetic disorders spanning a broad clinical spectrum (supplementary table S3), including conditions associated with *CSF1R* (MIM: 618476 and MIM: 221820), *SLC39A4* (MIM: 201100), *CLN8* (MIM: 600143), *ALDH4A1* (MIM: 239510), and *POLR1C* (MIM: 616494). In several cases, WES/WGS had identified potentially pathogenic or compound heterozygous variants, but the transcript-level consequences were unclear. These cases therefore exemplified scenarios in which targeted long-read RNA sequencing could provide orthogonal, mechanism-level evidence to refine variant interpretation, confirm a molecular diagnosis, or clarify the underlying disease mechanism. All samples were collected in PAXgene blood RNA tubes to support integration with clinically deployable workflows and minimise barriers to adoption.

To maximise translational relevance, we adopted a streamlined wet-lab protocol built from components already familiar within diagnostic laboratories. RNA extracted from peripheral blood was converted to cDNA using a low-input amplification workflow, and gene-specific primer sets were designed to enrich near-full-length transcripts for each candidate gene (see Figure 1A). Amplicons were barcoded using Oxford Nanopore Technologies (ONT) native barcoding system and pooled for long-read sequencing, allowing all six probands to be processed in a single run. This strategy enabled rapid turnaround while maintaining sufficient depth for transcript quantification. The complete laboratory protocol is publicly available via protocols.io (RAPID - RNA Analysis Pipeline for Integrated Diagnostics), reflecting its intentional design as a portable and reproducible workflow suited to diagnostic implementation.

**Figure 1.**
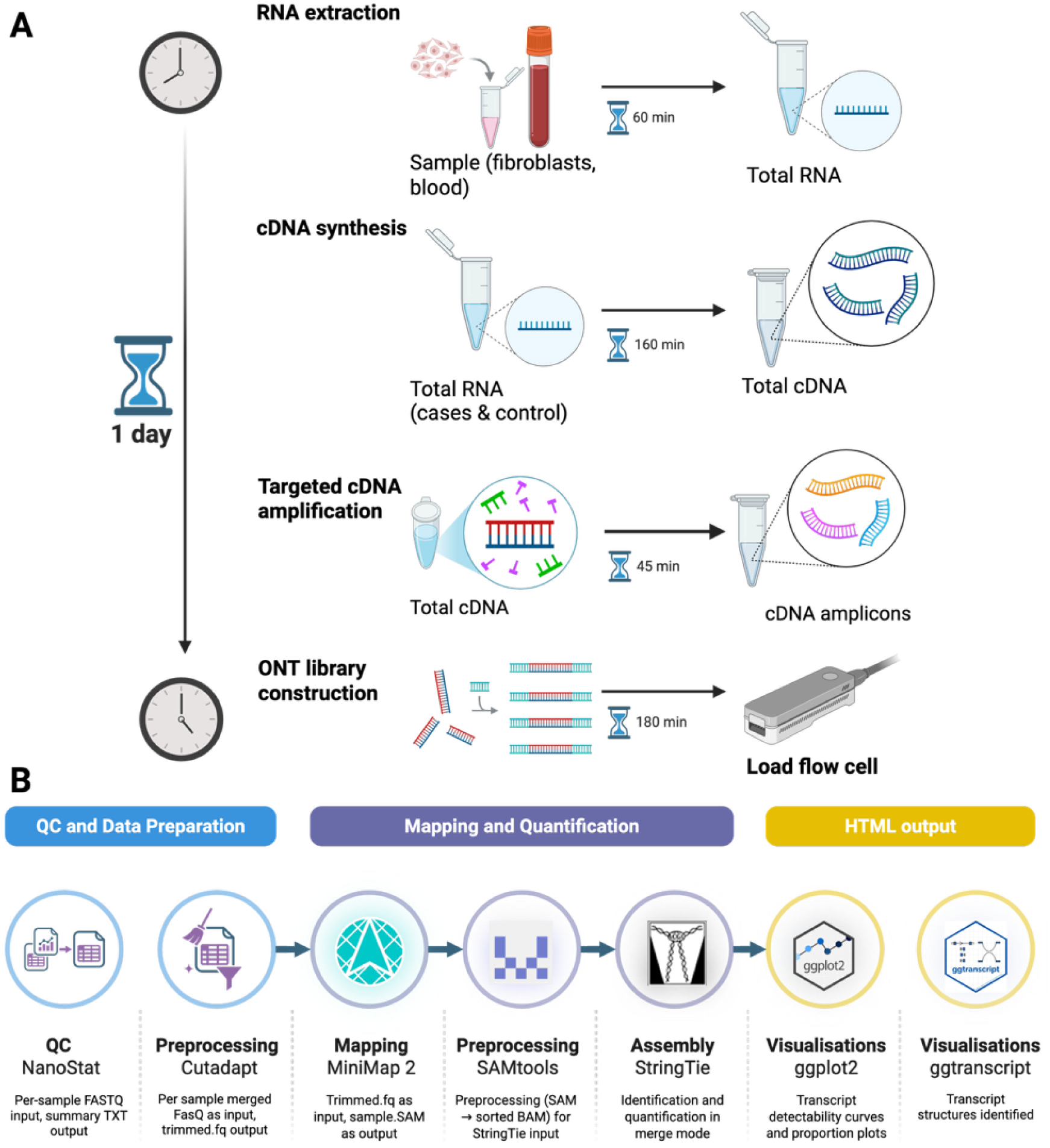
End-to-end wet and dry lab protocols: RAPID (RNA Analysis Pipeline for Integrated Diagnostics). (A) Wet lab methods; RNA extraction: Total RNA from patient peripheral blood was extracted using the PAXgene Blood RNA Kit (Qiagen) as per the manufacturer’s instructions. A commercially available pooled human blood RNA control (peripheral leukocytes; Takara Bio), comprising samples from 10 male and 3 female donors, was included as a reference. Total RNA was quantified using NanoDrop and Agilent TapeStation 4200 with the RNA ScreenTape analysis. **cDNA synthesis, primer design, and amplification:** Complementary DNA (cDNA) was synthesised using the NEBNext Single Cell/Low Input cDNA Synthesis & Amplification Module (New England Biolabs) according to the manufacturer’s instructions and a final amplification step of 11 PCR cycles. Primers were designed using NCBI Primer-BLAST against genome build GRCh38. Designs were verified against predicted products using the UCSC browser in-silico PCR tool and synthesised by Invitrogen; full sequences are listed in **supplementary table S1**. PCR amplification was performed using VeriFi HotStart Mix (PCR Biosystems) for 25 cycles according to the manufacturer’s instructions, see supplementary table S1 for further details of extension times. **Library preparation and sequencing:** The sequencing library was prepared from PCR amplicons using the Native Barcoding Kit 24 (v14 Oxford Nanopore Technologies) following the manufacturer’s protocol, Ligation sequencing amplicons - Native Barcoding Kit 24 (V14; SQK-NBD114.24). Libraries were sequenced on a MinION device with an R10 flow cell for 72 hours. Real-time base calling was performed in MinKNOW (v.24.11) using Dorado (v.0.9.0). **(B) Dry lab methods; Pre-processing:** Post-sequencing processing was conducted in R (v4.0.5) and the command line. Cutadapt (v5.0) was used to perform demultiplexing and adapter/primer trimming (per-amplicon adapters specified in a text file referenced in the config.yaml; minimum read length 200 nt unless noted). Read-level QC captured yield, read N50, and median Q-score. **Alignment:** Processed reads were mapped to the GRCh38 human reference genome using minimap2 (v2.28) with gene annotations from GENCODE (v.38) in splice-aware mode using platform-specific long-read sequencing presets (-ax splice). Secondary alignments were suppressed, and alignment was multi-threaded in order to improve specificity and increase alignment speed. **Transcript inference and modelling:** Long-read transcript models were inferred and collapsed with StringTie (v2.2.3) in merge mode, producing per-sample GTFs, a final merged GTF allowing consistent transcript naming and abundance estimates. Models were compared against GENCODE (v.47, GRCh38.p14, Ensembl release 113) and MANE-Select annotations to classify known versus novel transcript structures and to highlight differences in transcript structure when comparing to the MANE Select structure. **Quantification and case–control comparison:** For each gene of interest, the workflow outputs three key figures for interpretation. The first is a series of three line plots showing transcript detection rates in the case sample, in the control sample, and an overlay of case and control for direct comparison. The second is a bar plot showing the expression of each transcript in the case and control samples, expressed as a proportion of all transcripts generated from the locus in the given sample type. The third figure visualises transcript structures, aligned against the gene’s MANE-Select reference to highlight regions of relative gain or loss with ClinVar (release 2025-01-08) likely pathogenic and pathogenic variants in a track below to highlight regions sensitive to variation.

Computational analysis was carried out using a modular Snakemake^8^ workflow developed to operate end-to-end from raw FASTQ files. Reads underwent adapter trimming and demultiplexing, followed by long-read alignment and transcript model reconstruction to capture full-length isoforms (see Figure 1B). Case-control comparisons were generated using pooled reference samples sequenced in parallel, enabling proportional assessment of isoform usage and detection of aberrant or novel transcripts. The workflow produces clinician-facing HTML summaries and tabular outputs designed to support MDT interpretation. All tool configurations and dependencies are consolidated within a version-locked environment to ensure reproducibility across institutions. The workflow is publicly available at https://github.com/KylieMontgomery/RAPID, providing an openly accessible, clinically oriented pipeline for long-read RNA interpretation in as few as a single-sample, a capability not available in existing long-read diagnostic frameworks.

Analytical summaries were reviewed in multidisciplinary meetings with the relevant clinical teams. Interpretation followed three core principles: (i) identification of a novel transcript or splice junction present in the case but absent from controls and existing GENCODE v47 annotation; (ii) proportional reduction or absence of a transcript abundantly expressed in controls, consistent with splice disruption or exon skipping; and (iii) reduced transcript abundance consistent with nonsense-mediated decay in the presence of a pathogenic or exonic variant of uncertain significance. These transcript-level findings were then incorporated into clinical discussions to refine diagnosis, guide targeted DNA follow-up, or support variant reclassification. Further methodological detail, including tool evaluation (supplementary table S2), primer design (supplementary table S1) and sequencing metrics (supplementary table S4), is provided in the Supplementary material.

Applying this targeted long-read RNA-sequencing workflow to the six selected cases generated near-full-length transcript profiles of all candidate genes with sufficient depth for isoform-level interpretation. Using the interpretive framework defined above, namely the detection of novel transcript structures, loss or imbalance of expected isoforms, and evidence of nonsense-mediated decay, we identified four distinct modes through which RNA analysis contributed to diagnostic resolution (Table 1). In two cases, RNA-level evidence confirmed splice disruption and enabled reclassification of variants previously deemed uncertain. In a further two cases, transcript imbalance highlighted regions of interest for targeted DNA investigation. In one case, preserved splicing allowed confident exclusion of a candidate gene, while in another, complex splicing architecture limited interpretability but still provided actionable guidance by indicating the need for alternative approaches.

**Table 1.** Case interpretation based on predetermined criteria.

| Case | Novel transcript in case | Loss/reduction of transcript(s) in case | Reduced abundance consistent with NMD |
| --- | --- | --- | --- |
| P1 | ✓ | ✓ | ✓ |
| P2 | – | ✓ | – |
| P3 | – | ✓ | – |
| P4 | ✓ | ✓ | N/A |
| P5 | – | – | – |
| P6 | – | ✓ | – |

For patients P1 and P2, long-read RNA sequencing provided direct, full-length evidence of splice disruption in *CSF1R*, enabling definitive molecular diagnoses. Patient P1, presenting with childhood-onset neurodevelopmental features, carried a pathogenic variant (NM_001288705.3: c.840del (p.Ser281AlafsTer16)) together with a splice-region variant of uncertain significance (NC_000005.10(NM_001288705.3): c.592+5A>G).

Long-read analysis identified a novel exon-3 skipping transcript exclusive to the case, absent from pooled controls and GTEx, and accounting for 65% of detected transcripts. The canonical isoform was markedly reduced, consistent with nonsense-mediated decay, and splice-aware alignment confirmed a clean exon 2 to exon 4 junction. These findings provided clear functional validation that the c.592+5A>G variant disrupts splicing, supporting its reclassification as likely pathogenic and establishing a diagnosis of *CSF1R*-related brain abnormalities, neurodegeneration, and dysosteosclerosis (BANDDOS, MIM:618476) (see Figure 2 A-C). Patient P2, an adult male with a progressive leukoencephalopathy, harboured a single heterozygous splice-region variant (NC_000005.10(NM_001288705.3):c.2763+4_2763+7del). RNA analysis similarly revealed a novel exon-20 skipping transcript comprising 63% of all transcripts and absent from relevant control datasets, providing orthogonal confirmation of splice disruption and supporting reclassification of the variant as likely pathogenic, consistent with a diagnosis of CSF1R-relatedadult-onset leukoencephalopathy with axonal spheroids and pigmented glia ( ALSP, MIM:221820)(see supplementary figure S1).

**Figure 2.**
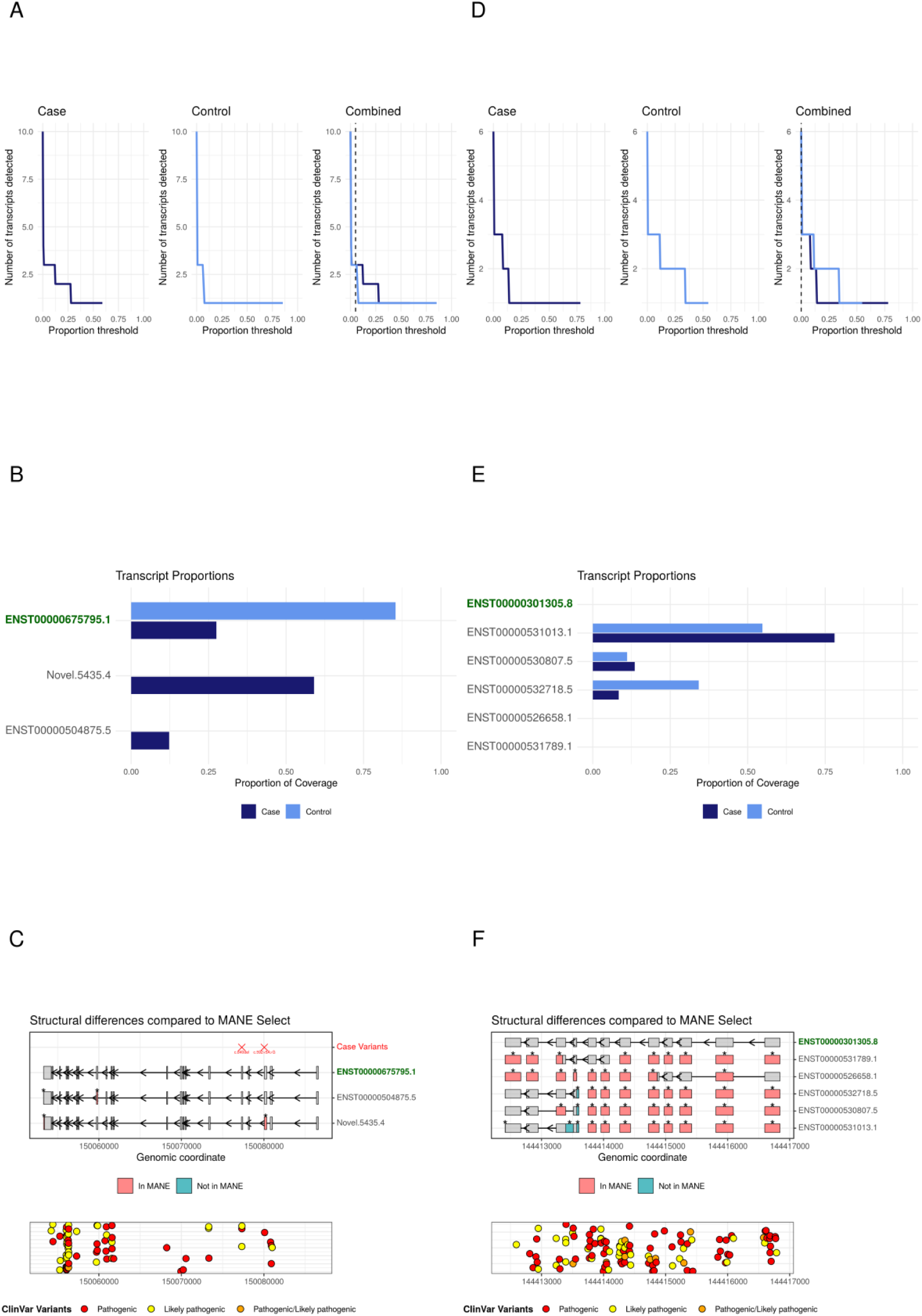
Cases P1 and P6 results. **(A, D)** Transcript detectability plots A clear shift in transcript detectability was observed between the case and pooled-control samples, where plateau regions were evident, indicative of multiple transcript populations reliably captured at increasing thresholds, as such a threshold of 5% was deemed appropriate. **(B, E) Proportions of each transcript detected over 5%** In plot C, a novel *CSF1R* transcript was identified exclusively in the case sample, absent from both pooled controls and GTEx annotations, and this structure accounted for 65% of all transcripts in the proband. In plot D, the case shows a single dominant transcript, the MANE-Select transcript (ENST00000642195.1), whereas in the pooled control, five unique transcripts were observed. This indicated a loss of transcript diversity. **(C, F) Transcript structure plots above the determined threshold (5%).** In figure E, the novel structure showed exon-3 skipping relative to the MANE-Select transcript (ENST00000675795.1), and this encodes part of the immunoglobulin-like domain essential for receptor signalling, suggesting functional disruption. In figure F, with the complexity of the *POLR1C* architecture, a candidate region for a causative variant(s) is challenging to ascertain.

In patients P3 and P4, long-read RNA sequencing did not identify a single definitive splice alteration but revealed systematic transcript-level disequilibrium that localised candidate regions for further DNA analysis. In patient P3, presenting with features consistent with acrodermatitis enteropathica but lacking an exonic *SLC39A4* variant, the case sample showed a single dominant transcript accounting for 78% of all reads. In contrast, the pooled controls displayed no dominant isoform and instead expressed five transcripts at broadly similar proportions, including the dominant transcript identified in the case. All detected structures in the pooled-control converged on a shared region within intron 9 of the MANE-Select transcript, pointing to a potential deep-intronic variant driving abnormal transcription in the case (see Figure 2 D-F).

These observations informed MDT consensus to undertake targeted sequencing of this region. Patient P4, a child with a progressive neurodegenerative phenotype and ultrastructural features in blood film suggestive of a variant form of neuronal ceroid lipofuscinosis, but only a single heterozygous *CLN8* variant identified by trio WGS, (NM_018941.4:c.67_76delinsT (p.Ile23_Thr26delinsSer)), demonstrated numerous transcript shifts across a highly spliced region with no dominant isoform in either case or control. This disequilibrium raised suspicion for a splice-altering variant not prioritised by WES or WGS analysis. Re-evaluation is now focused on the donor region of intron 2 of the MANE-Select transcript, which aligns with the observed transcript perturbations and illustrates how long-read RNA data can direct second-allele searches when DNA-based testing is inconclusive (see supplementary figure S2).

Patient P5 presented with developmental delay, seizures and hyperprolinaemia, however transcript-level data enabled confident exclusion of *ALDH4A1* as a candidate gene. The proband and pooled control exhibited a highly similar transcript profile, with a preserved MANE-Select isoform and a low-abundance novel transcript present in both samples. The absence of any case-specific splicing alteration supported removal of *ALDH4A1* from diagnostic consideration, and the clinical team subsequently identified a molecular diagnosis involving an alternative gene, prioritized under review due to an evolving phenotype (see Figure 3 A-B). Patient P6, by contrast, illustrated the limitations posed by inherently complex transcript architecture. The proband displayed a congenital, multisystem phenotype of mid-face and mandibular hypoplasia reminiscent of Treacher Collins syndrome and white matter changes on brain MRI scan. suggestive of a disorder involving *POLR1C*. Long-read RNA sequencing revealed no dominant transcript in either case or control. Instead, the case showed reduced transcript diversity relative to pooled controls, without a discrete region of interest that could be prioritised for variant exploration. These findings highlighted the biological complexity of *POLR1C* splicing and guided MDT recommendations for renewed variant review of WGS data, recognising that certain structural, deep-intronic, or regulatory variants may require alternative or enhanced DNA sequencing strategies beyond RNA-based approaches (see Figure 3 C-E).

**Figure 3.**
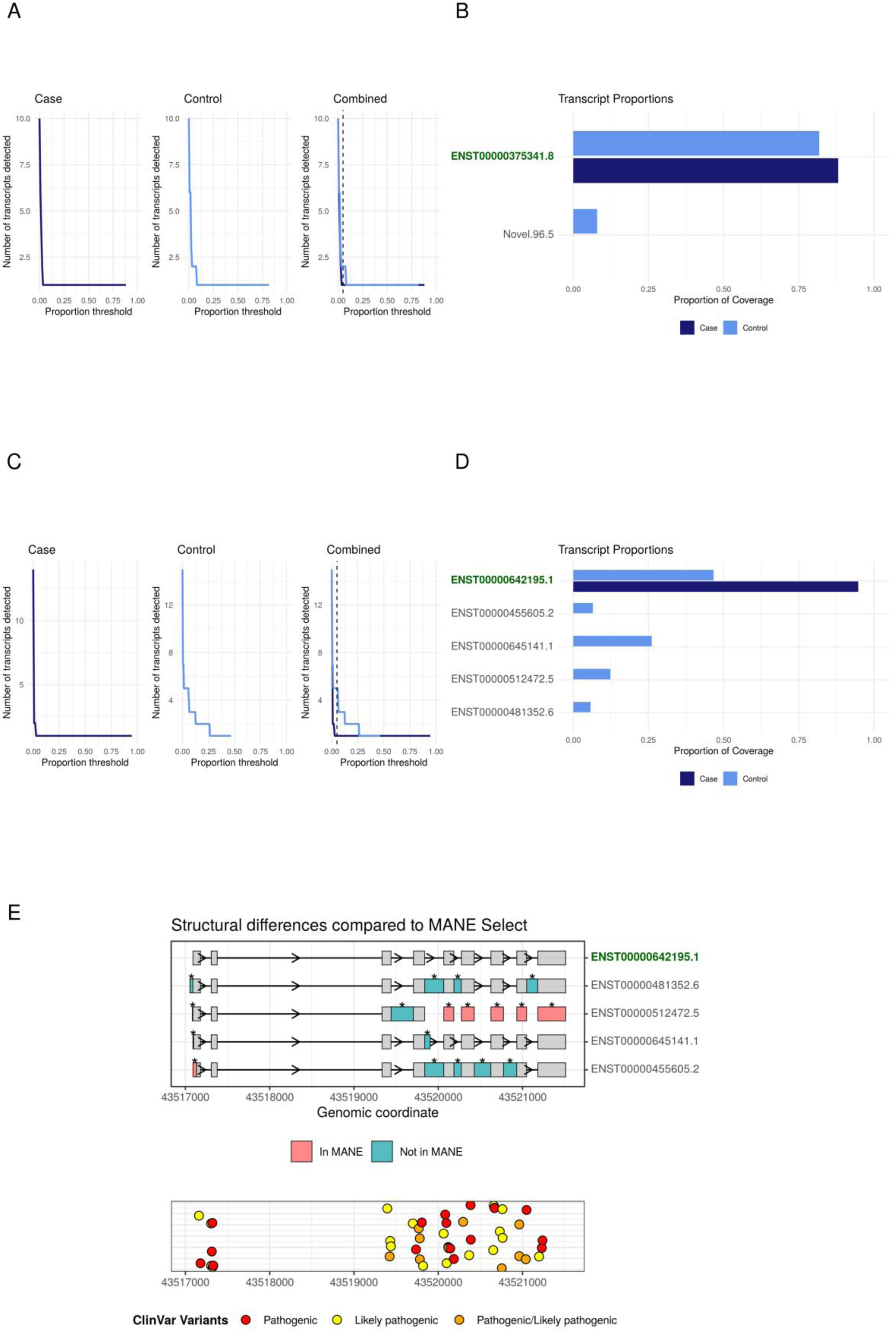
Cases P2 and P5 results. **(A, C)** In figure A, there is almost no difference in transcript detectability between case and control. In figure B, a clear shift in transcript detectability was observed between the case and pooled-control samples, where plateau regions were evident, indicative of multiple transcript populations reliably captured at increasing thresholds, as such a threshold of 5% was deemed appropriate. **(B, D) Proportions of each transcript detected over 5%** In plot C, a novel *ALDH4A1* transcript was identified exclusively in the control sample however, st 8% it is difficult to ascertain if this is insufficient sequencing depth, natural variation or something more meaninful. In plot D, case and controls show that three unique transcripts were observed. There is a shift in proportions which only when considering the transcript structures does a region in common appear. **(E) Transcript structure plot above the determined threshold (5%).** In figure E, closer inspection reveals a region in common across the three detected structures aligning to the MANE-Select (ENST00000301305.8) intron 9, indicating a potential prioritisation of an alternative start site receptor signalling, suggesting functional disruption. Intronic and regions are not covered and coverage is often reduced at junction sites by targeted-exome panels as such, orthogonal DNA testing (targeted sequencing of the region of interest) is underway to resolve a potential causative variant.

The RAPID workflow was developed to address directly the barriers that have limited the clinical use of long-read RNA sequencing. Targeted long-read RNA sequencing offers a powerful way to resolve transcript structure and splicing, yet most published approaches remain difficult to implement in diagnostic settings because they rely on whole-transcriptome sequencing, complex analysis pipelines, and extensive reference cohorts. RAPID overcomes these challenges by using targeted enrichment, pooled controls, and a portable analysis pipeline that supports single-sample interpretation, thereby removing key logistical barriers and enabling routine diagnostic deployment. In this pilot study, the workflow generated informative transcript profiles for all probands and demonstrated that long-read RNA sequencing can be incorporated into diagnostic pathways in a reliable and cost-effective manner.

The clinical impact of this approach was evident across the cohort. In two cases, RAPID provided direct transcript-level evidence of splice disruption that supported the reclassification of variants and delivered confirmed molecular diagnoses. In two further cases, systematic perturbations in transcript proportions highlighted regions within intronic or regulatory sequence that required targeted DNA investigation, thereby refining the search for a second allele in suspected recessive disease. Another case demonstrated that preserved splicing can support the exclusion of a candidate gene, helping to focus diagnostic interpretation. The final case illustrated that some genes with highly complex transcript structure may remain challenging to resolve fully, yet the RNA findings still informed subsequent testing strategies. Together, these outcomes demonstrate that long-read RNA data can deliver decisive mechanistic insights across diverse diagnostic scenarios, including variant confirmation, second-allele localisation, gene exclusion, and strategic triage for further testing. In all scenarios, the ability to obtain clear, mechanistically informative results within clinically relevant timeframes underscores the value of integrating long-read RNA evidence into routine diagnostic practice.

Although the workflow performed well across all cases, several limitations should be recognised. Interpretation relies on adequate expression of the relevant gene in peripheral blood, and alternative tissues may be needed for genes expressed at low levels. Amplification efficiency is influenced by primer design, and very large or GC-rich amplicons can be difficult to capture. Genes with extensive isoform diversity or repetitive architecture can also limit the precision of transcript assignment, even with full-length reads. Long-read sequencing provides reliable isoform-level information but is not optimised for precise nucleotide-level variant calling, so complementary DNA-based testing may still be required when RNA findings are suggestive but not diagnostic. Continued updates to basecalling models and reference annotations also require ongoing evaluation, though the modular and fully version-locked architecture of RAPID is specifically designed to accommodate such evolution without disrupting clinical reproducibility.

In summary, RAPID combines the biological resolution of long-read RNA sequencing with a workflow that is feasible for diagnostic laboratories. By integrating targeted enrichment, reproducible analysis, and streamlined interpretive outputs, this approach provides a scalable and cost-effective method for assessing splicing abnormalities in rare disease. As a clinically deployable long-read RNA framework, RAPID fills a key gap between discovery-oriented sequencing technologies and the practical requirements of diagnostic genomics. With appropriate tissue selection and continued refinement, long-read RNA sequencing is well positioned to enhance functional interpretation and improve diagnostic outcomes in clinical genomics.

## Data Availability

The data underlying this study consist of human participant data and include potentially identifying clinical and molecular information. Due to the conditions of ethical approval and the informed consent provided by participants the raw data cannot be made publicly available.
De-identified or aggregated data supporting the findings of this study may be made available from the corresponding author upon reasonable request subject to approval by the relevant ethics committee and in accordance with applicable data protection regulations.

## Notes

### Competing Interest Statement

The authors have declared no competing interest.

### Clinical Protocols

https://github.com/KylieMontgomery/RAPID

https://www.protocols.io/view/rapid-rna-analysis-pipeline-for-integrated-diagnos-g37hbyrj7

### Funding Statement

This study was funded by Great Ormond Street Hospital Childrens Charity and Sparks National Funding Call (reference: V4021)

### Author Declarations

The ethics committee of the UCL GOS Institute of Child Health, London (13/LO/0168) and the UCL Queen Square Institute of Neurology, London (22/NE/0080) gave ethical approval for this work.

## References

1. Boycott KM, Ardigó D. Addressing challenges in the diagnosis and treatment of rare genetic diseases. Nat Rev Drug Discov.Nature Publishing Group. 2018;17(3):151–152. doi:10.1038/nrd.2017.246

2. The Lancet Global Health. The landscape for rare diseases in 2024. Lancet Glob Health.Elsevier Ltd. 2024;12(3):e341. doi:10.1016/S2214-109X(24)00056-1

3. Wai HA, Lord J, Lyon M, et al. Blood RNA analysis can increase clinical diagnostic rate and resolve variants of uncertain significance. Genetics in Medicine. Published online 2020:22. doi:10.1038/s41436

4. Lord J, Baralle D. Splicing in the Diagnosis of Rare Disease: Advances and Challenges. Front Genet.Frontiers Media S.A. 2021;12. doi:10.3389/fgene.2021.689892

5. Cummings BB, Marshall JL, Tukiainen T, et al. Improving genetic diagnosis in Mendelian disease with transcriptome sequencing. Sci Transl Med. 2017;9(386). doi:10.1126/scitranslmed.aal5209

6. Kremer LS, Bader DM, Mertes C, et al. Genetic diagnosis of Mendelian disorders via RNA sequencing. Nat Commun. 2017;8. doi:10.1038/ncomms15824

7. Yépez VA, Mertes C, Müller MF, et al. Detection of aberrant gene expression events in RNA sequencing data. Nat Protoc. 2021;16(2):1276–1296. doi:10.1038/s41596-020-00462-5

8. Köster J, Rahmann S. Snakemake-a scalable bioinformatics workflow engine. Bioinformatics. 2012;28(19):2520–2522. doi:10.1093/bioinformatics/bts480

9. Martin M. Cutadapt removes adapter sequences from high-throughput sequencing reads. EMBnet J. 17(1):10–12. doi:10.14806/ej.17.1.200

10. Pertea M, Pertea GM, Antonescu CM, Chang TC, Mendell JT, Salzberg SL. StringTie enables improved reconstruction of a transcriptome from RNA-seq reads. Nat Biotechnol. 2015;33(3):290–295. doi:10.1038/nbt.3122

11. Wickham Hadley. Ggplot2: Elegant Graphics for Data Analysis. Springer; 2009.

12. Gustavsson EK. Ggtranscript: An R Package for the Visualization and Interpretation of Transcript Isoforms Using Ggplot2. Springer; 2022. doi:10.1093/bioinformatics/btac409

13. Oxford Nanopore Technologies. Dorado (v0.5.1). 2023. https://github.com/nanoporetech/dorado

