## supplementary figure S1 for "RAPID: A Targeted Long-Read RNA Workflow for Functional Resolution of Splicing Variants in Rare Disease"

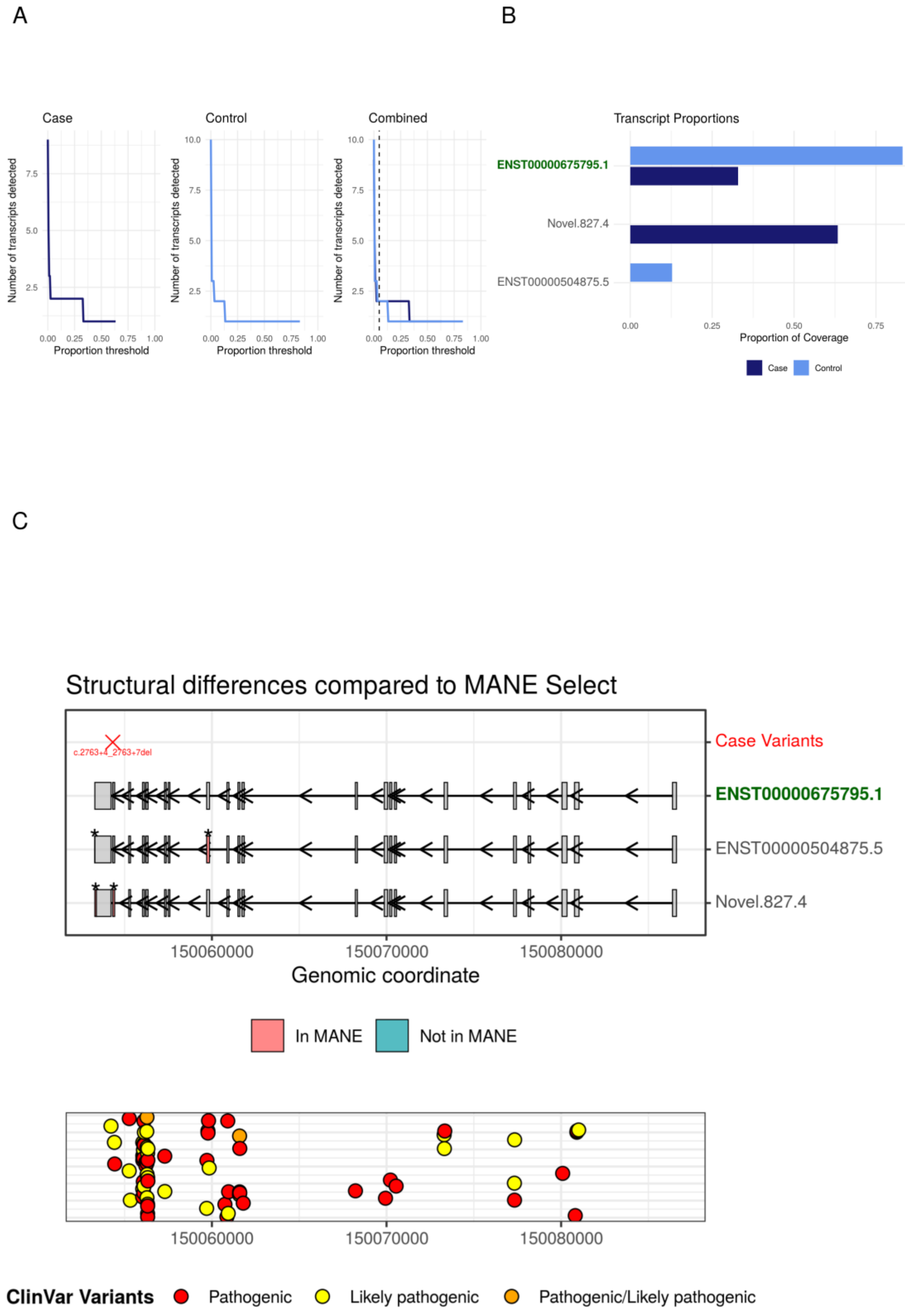

A
B
Case
Control
Combined
Number of transcripts detected
Proportion threshold
Transcript Proportions
ENST00000675795.1
Novel.827.4
ENST00000504875.5
Proportion of Coverage
Case
Control
C
Structural differences compared to MANE Select
c.2763+4_2763+7del
Case Variants
ENST00000675795.1
ENST00000504875.5
Novel.827.4
150060000
150070000
150080000
Genomic coordinate
In MANE
Not in MANE
ClinVar Variants
Pathogenic
Likely pathogenic
Pathogenic/Likely pathogenic

**Supplementary Figure S1. Patient P2 (*CSF1R*) (A)** Identification of a suitable threshold was achieved with the utilisation of the transcript detectability plots generated by the RAPID pipeline. There are multiple plateau regions seen in both case and pooled-control in order to capture these events, a threshold of 5% was chosen. **(B)** A novel *CSF1R* transcript was identified exclusively in the case sample, absent from both pooled controls and GTEx annotations, and accounted for 63% of all transcripts in the proband. (C) The novel *CSF1R* transcript structure showed exon-20 skipping relative to the MANE-Select transcript (ENST00000675795.1) and accounted for 63\% of all transcripts in the proband. Exon 20 encodes part of the catalytic domain essential for receptor signalling, suggesting functional disruption.
