## supplementary figure S2 for "RAPID: A Targeted Long-Read RNA Workflow for Functional Resolution of Splicing Variants in Rare Disease"

A

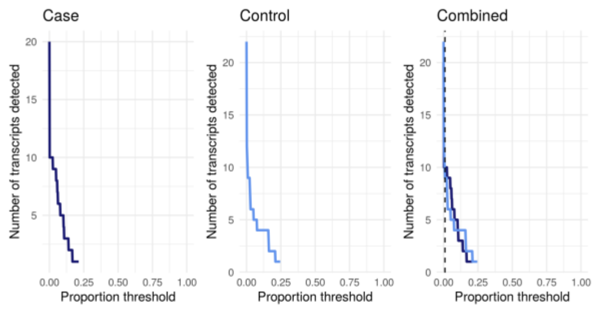
Case
Control
Combined
Number of transcripts detected
Proportion threshold
0.00
0.25
0.50
0.75
1.00

B

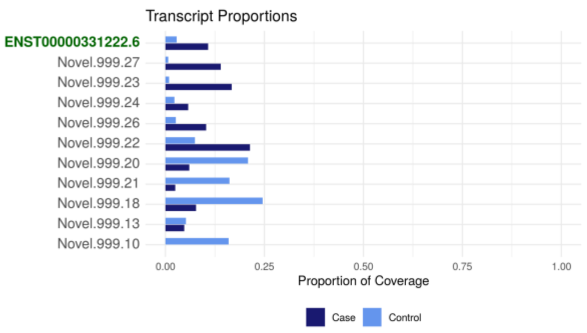
Transcript Proportions
ENST00000331222.6
Novel.999.27
Novel.999.23
Novel.999.24
Novel.999.26
Novel.999.22
Novel.999.20
Novel.999.21
Novel.999.18
Novel.999.13
Novel.999.10
Proportion of Coverage
Case
Control

C

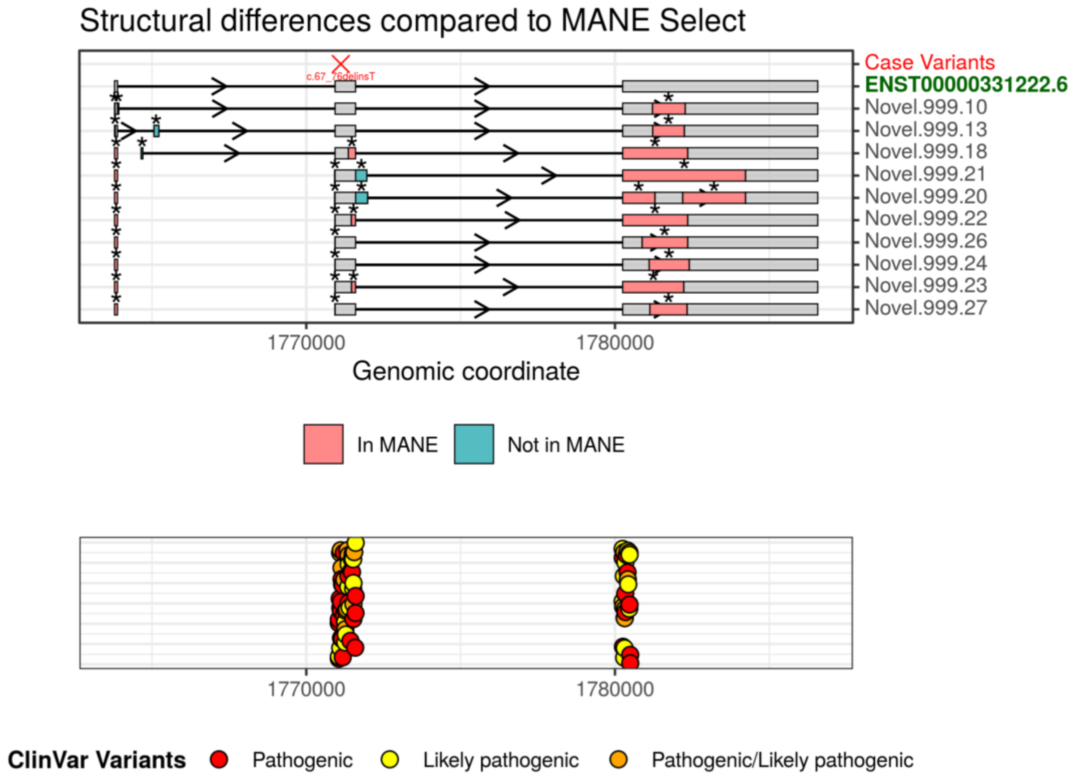
Structural differences compared to MANE Select
c.67_75delinsT
Case Variants
ENST00000331222.6
Novel.999.10
Novel.999.13
Novel.999.18
Novel.999.21
Novel.999.20
Novel.999.22
Novel.999.26
Novel.999.24
Novel.999.23
Novel.999.27
1770000
1780000
Genomic coordinate
In MANE
Not in MANE
ClinVar Variants
Pathogenic
Likely pathogenic
Pathogenic/Likely pathogenic

**Supplementary Figure S2. Patient P4 (*CLN8*). (A)** Identification of a suitable threshold was achieved with the utilisation of the transcript detectability plots generated by the RAPID pipeline. There are numerous plateau regions and no single predominant transcript seen in both case and pooled-control. In order to capture these events, a reduced threshold of 1% was chosen. **(B)** Inspecting the transcript proportion barplot reveals a high number of novel transcripts identified in both the case and pooled-control samples, with no single transcript exceeding 25% of the total detected. With deviations from the control evident, it was important to assess the transcript structures to ascertain if this was connected to the previously identified variant or an indicator of a candidate for investigating deep intronic variants. **(C)** Reciprocal shifts across several transcripts within the gene's highly spliced region were identified. This transcript disequilibrium raises suspicion for a splice-altering variant. With a secondary pathogenic variant not identified by trio-WGS, a candidate variant may be a rare synonymous change. Variant re-evaluation prioritising the donor region of MANE-Select (ENST00000331222.6) intron 2 is in progress.
