## supplementary methods for "RAPID: A Targeted Long-Read RNA Workflow for Functional Resolution of Splicing Variants in Rare Disease"

**Ethics:**

**Paediatric cohort**

Ethical and health research authority (HRA) approval was granted for collection and analysis of samples from patients under the Study of Inherited Metabolic Diseases (SIMD):
IRAS Project ID: 95005
REC Reference: 13/LO/0168
NRES Committee: Bloomsbury, London
This project is also listed on the NIHR Clinical Research Network Portfolio.

In addition, patients and families participating in the 100,000 Genomes project were recruited under NHS England (NHSE) ethics approval, which permits researchers to recontact participants up to 4 times per year for follow-up data or sample requests. These activities were conducted under the framework of the Genomics England Clinical Interpretation Partnerships (GeCIPs), of which Professor Ryten and myself are members.

**Adult cohort**

Under local ethics approval peripheral blood was collected in PAXgene RNA tubes from individuals with suspected neurological disorders as part of an observational study with international collaborators, to identify disease genes, genetic and COVID-19 risk factors, disease biomarkers and fundamental genetic mechanisms (International Genomics Collaboration - IGC).
IRAS Project ID: 310045
REC Reference: 22/NE/0080
NRES Committee: Bloomsbury, London

Consent and blood sample collection was performed by the appropriate clinical team. All samples were stored at -80ºC and pseudo anonymised before accepted by the Ryten group.
