## supplementary table S1 for "RAPID: A Targeted Long-Read RNA Workflow for Functional Resolution of Splicing Variants in Rare Disease"

| Gene | Forward primer | Reverse primer | PCR extension time |
|---|---|---|---|
| *CSF1R* | GGAGAGGAACGTGTGTCCAG | TCTAGCCCAGAATGACGGGA | 2 min |
| *SLC39A4* | GACTGAGCCCAGGGGACTTC | GTTGGGACTGGGGCCTCTAT | 1 sec |
| *CLN8* | GGCAGCCCAGATTGAAGATG | TGGGTGCGTCCTGTCTTTAC | 3 min |
| *ALDH4A1* | GAACAGCCCCGCTTCTAACC | ATCCCATTGCCACCTCACAG | 1.5 min |
| *POLR1C* | GTTTGGGGTTCGCAATGTCC | GGGCTCTCTGTTCAGCAGTC | 30 sec |
