## supplementary table S2 for "RAPID: A Targeted Long-Read RNA Workflow for Functional Resolution of Splicing Variants in Rare Disease"

| Tool | Platform compatibility | Native barcoding constraints | Transcript discovery | Maintenance & longevity | Other |
|---|---|---|---|---|---|
| Deepbinner | ✗ | ✓ | – | ✗ | Deprecated since 2021 |
| Lima | ✗ | ✓ | – | ✗ | For use with PacBio Hi-Fi reads |
| Porechop | ✗ | ✓ | – | ✗ | For use with ONT reads, unsupported since 2018 |
| Pychopper | ✗ | ✗ | – | ✓ | For use with rapid ONT barcoding |
| Cutadapt | ✓ | ✓ | – | ✓ | – |
| StringTie | ✓ | – | ✓ | ✓ | – |
| FLAIR | ✓ | – | ✓ | ✓ | – |
| Mandalorian | ✗ | – | ✓ | ✓ | Tested for use with Lima output (PacBio) |
| Bambu | ✓ | – | ✓ | ✓ | Biased toward existing annotation |
| TALON | ✓ | – | ✗ | ✓ | Python package |
| FREDDIE | ✓ | – | ✓ | ✓ | Must run gffcompare for quantification |
| Isoquant | ✓ | – | ✓ | ✓ | Expects poly-A tails |
| FLAMES | ✓ | – | ✓ | ✓ | Requires a fix using BAMBU |
| SQANTI3 | ✓ | – | ✓ | ✓ | References a deprecated file |
| ggtranscript | – | – | – | ✓ | – |
| IsoformSwitchAnalyzeR | – | – | – | ✓ | Computationally excessive for project requirements |
| Genoverse | – | – | – | ✓ | Complex installation and extensive customisation for project requirements |
