## supplementary table S3 for "RAPID: A Targeted Long-Read RNA Workflow for Functional Resolution of Splicing Variants in Rare Disease"

| Patient | Onset | Key clinical features | Candidate gene | DNA variant(s) | Zygosity | Prior testing | Diagnosis (provisional) |
|---|---|---|---|---|---|---|---|
| P1 | Childhood | Optic atrophy; sagittal craniosynostosis; metaphyseal dysplasia; intracranial calcification; developmental delay | *CSF1R* | NM_001288705.3:c.840del (p.Ser281AlafsTer16)/ NC_000005.10(NM_001288705.3):c.592+5A>G | Comp Het | WGS | Brain abnormalities, neurodegeneration, and dysosteosclerosis (BANDDOS (MIM: 618476)) |
| P2 | Adulthood | Gait apraxia; dysexecutive change; WM disease (CC involvement with cerebral atrophy) | *CSF1R* | NM_001288705.3:c.2763+4_2763+7del | Het | WGS | Adult-onset leukoencephalopathy with axonal spheroids and pigmented glia (ALSP (MIM: 221820)) |
| P3 | Infancy | Lethargy and oliguria. Identified narrow-complex tachycardia; subsequently diagnosed with Wolff–Parkinson–White syndrome. Severe eczema with persistent anal fissures; low zinc levels identified. Diagnosed with acrodermatitis enteropathica. Recurrent coughs and upper respiratory tract infections | *SLC39A4* | – | – | WES R332 panel | Zinc metabolism disorder, Acrodermatitis enteropathica? |
| P4 | Childhood | Progressive neurodegenerative phenotype and ultrastructural features in blood film suggestive of a variant form of neuronal ceroid lipofuscinosis | *CLN8* | NM_018941.4:c.67_76delinsT (p.Ile23_Thr26delinsSer) | Het | Extensive metabolic testing<br>Array CGH – normal<br>SCA1, 2, 3 & 6 genes – negative<br>Mitochondrial genome – negative<br>Fragile-X - negative<br>WGS; R231 panel | Neuronal ceroid lipofuscinosis-8 (MIM: 600143) |
| P5 | Infancy | Hydronephrosis detected in pregnancy, Dev delay, learning difficulty, autism, Seizures, EEG showed left focal changes, diagnosis of mild hyperprolinaemia, grey matter subependymal heterotopia | *ALDH4A1* | – | – | WGS | Hyperprolinemia type II (MIM: 239510) |
| P6 | Infancy | IUGR; short stature; distinctive facies; progressive leukodystrophy; bilateral SNHL; bilateral cryptorchidism | *POLR1C* | – | – | Trio WGS-normal; Array-normal; SRS-normal | – |
