## supplementary table S4 for "RAPID: A Targeted Long-Read RNA Workflow for Functional Resolution of Splicing Variants in Rare Disease"

| Case | Target | Expected amplicon length | N50 | Total reads |
|---|---|---|---|---|
| P1 | *CSF1R* | 3,654 | 3,591 | 160,000 |
| P2 | *CSF1R* | 3,654 | 3,612 | 106,210 |
| P3 | *SLC39A4* | 426 | 546 | 194,122 |
| P4 | *CLN8* | 6,272 | 2,371 | 106,210 |
| P5 | *ALDH4A1* | 3,093 | 3,146 | 648,000 |
| P6 | *POLR1C* | 1,064 | 1,179 | 409,802 |
